# Algorithmic identification of treatment-emergent adverse events from clinical notes using large language models: a pilot study in inflammatory bowel disease

**DOI:** 10.1101/2023.09.06.23295149

**Authors:** Anna L Silverman, Madhumita Sushil, Balu Bhasuran, Dana Ludwig, James Buchanan, Rebecca Racz, Mahalakshmi Parakala, Samer El-Kamary, Ohenewaa Ahima, Artur Belov, Lauren Choi, Monisha Billings, Yan Li, Nadia Habal, Qi Liu, Jawahar Tiwari, Atul J Butte, Vivek A Rudrapatna

**Author notes:** **Corresponding Author:** Vivek A. Rudrapatna, MD, PhD, Assistant Professor of Medicine, University of California, San Francisco Bakar Institute, Box 2993, 490 Illinois Street, Floor 2, San Francisco, CA 94143. These authors contributed equally to this work and share first authorship (Anna L Silverman, Madhumita Sushil, Balu Bhasuran). This work was done while at the FDA. Current affiliation University of Maryland School of Medicine, Baltimore, MD, USA.

## Abstract

**Background and Aims:** Outpatient clinical notes are a rich source of information regarding drug safety. However, data in these notes are currently underutilized for pharmacovigilance due to methodological limitations in text mining. Large language models (LLM) like BERT have shown progress in a range of natural language processing tasks but have not yet been evaluated on adverse event detection.

**Methods:** We adapted a new clinical LLM, UCSF BERT, to identify serious adverse events (SAEs) occurring after treatment with a non-steroid immunosuppressant for inflammatory bowel disease (IBD). We compared this model to other language models that have previously been applied to AE detection.

**Results:** We annotated 928 outpatient IBD notes corresponding to 928 individual IBD patients for all SAE-associated hospitalizations occurring after treatment with a non-steroid immunosuppressant. These notes contained 703 SAEs in total, the most common of which was failure of intended efficacy. Out of 8 candidate models, UCSF BERT achieved the highest numerical performance on identifying drug-SAE pairs from this corpus (accuracy 88-92%, macro F1 61-68%), with 5-10% greater accuracy than previously published models. UCSF BERT was significantly superior at identifying hospitalization events emergent to medication use (p < 0.01).

**Conclusions:** LLMs like UCSF BERT achieve numerically superior accuracy on the challenging task of SAE detection from clinical notes compared to prior methods. Future work is needed to adapt this methodology to improve model performance and evaluation using multi-center data and newer architectures like GPT. Our findings support the potential value of using large language models to enhance pharmacovigilance.

## Introduction

The accurate detection of treatment-emergent adverse events (AEs) is critical to ensure that clinicians and patients can make well-informed treatment decisions that balance risks with benefits. This is particularly true of non-steroid immunosuppressants which are commonly needed long-term for the treatment of inflammatory bowel diseases (IBD).

Existing approaches for AE surveillance may involve prospective registry studies, spontaneous postmarketing reporting (e.g., the Food and Drug Administration’s AE Reporting System [FAERS])^1^, literature searches, and/or analyses of the structured data from claims and electronic health records databases^2,3^. These approaches have provided important data on the postmarket safety of medications but are limited by expense, small numbers, under/over-reporting^4,5^, missing data, limitations in inferring causality, and suboptimal sensitivity and specificity. Clinical notes are a rich source of AE data because treating clinicians often document actions in response to potential AEs, including treatment discontinuation and hospitalization. However, these notes have been underutilized for surveillance due to methodological limitations in effective text mining.

Recent years have seen impressive advances in natural language processing following the release of the large language model known as BERT (Bidirectional Encoder Representations from Transformers)^6^. However, its adaptation to domain-specific arenas like medical language has been limited, in part due to the unavailability of safe platforms for processing this protected health information until recently. In prior work, our group of academic researchers has developed a new BERT model specifically designed to interpret clinical text as typically documented in electronic health records (EHR) systems^7^. This model, UCSF BERT, was trained on 75 million clinical notes documented across a range of specialties over the last 10 years at the University of California, San Francisco (UCSF). Evaluations of UCSF BERT on several general benchmarks show that it performs as well as or better than other comparable BERT models not specifically trained from scratch using a diverse corpus of notes derived from EHRs^7^. However, these prior evaluations were general tasks and are limited by the quality of currently available, publicly benchmarked tasks.

An open question motivating this study was whether the BERT model could help automate specific tasks of established clinical importance, particularly one as challenging as AE detection. Many aspects make the task of AE detection from clinical notes particularly difficult. These include the length of typical clinical notes, the need to infer relationships between medications and documented AEs, to encode AEs in a standardized way, and to overcome inherent vagueness in the documentation of clinical notes. Some current examples of automated AE detection come from the National Natural Language Processing Clinical Challenges (N2C2)^8^ adverse event detection challenge, a nationwide clinical data science competition that was held in 2018. Models from this competition were evaluated on highly simplified benchmark tasks that do not reflect the typical patterns of clinical documentation such as short snippets of notes rather than full length notes. Notably, none of the candidate models from the competition were large language models as it was held prior to the wide-spread adoption of large language models.

We hypothesized that adaptations of the UCSF BERT, a large language model, would outperform previously published methods on multiple tasks related to AE detection, due in large part to its prior training on a large volume of EHR notes. In this pilot study, we trained UCSF BERT to identify hospitalization-associated serious adverse events (SAEs) from notes written in the outpatient IBD clinic, and we compared its performance to a range of baselines including previously published models.

## Methods

### Ethics

This single-center study of natural language processing algorithms for adverse event detection was approved by the UCSF Institutional Review Board (#18-24588).

### Target of prediction

The target of prediction was treatment-emergent, outpatient SAEs requiring hospitalization with exposure to non-steroid immunosuppressant drugs for IBD, as documented in the outpatient gastroenterology clinic notes at UCSF. The candidate list of drugs included all biologics and small-molecule medications, except steroids, that were approved by the FDA for the treatment of ulcerative colitis or Crohn’s disease as of 2020, as well as off-label medications that are occasionally used to treat these conditions. A complete list of included medications can be found in the supplemental materials. Although the FDA’s definition of SAEs includes multiple categories^9^, we limited our scope to only SAEs associated with a hospitalization event, as these are more likely to be well-documented in clinical notes due to their clinical importance. We defined treatment-emergent as an SAE that occurred while the patient was actively receiving scheduled doses of a given medication, having been absent pre-treatment. For example, if a patient was hospitalized for pneumonia 6 weeks after receiving an infusion that was prescribed to be given every 8 weeks, the hospitalization event would be considered a treatment-emergent SAE. Once the clinical decision to discontinue a given treatment plan was documented, subsequent hospitalization events were no longer considered treatment-emergent SAEs. Worsening of previously existing conditions that prompted hospitalization were included in line with internationally used guidelines on AE reporting^10^. A definitive assessment of potentially causal relationships between treatments and SAEs was beyond the planned scope of this analysis.

### Document identification strategy

To identify the target notes for this study, we used a deidentified research database consisting of structured EHR data at UCSF as well as clinical notes that had been subjected to automated redaction of protected health information^11^. We queried the database to identify all notes associated with the gastroenterology department and an IBD diagnosis code (ICD-9 555/556; ICD-10 K50/K51). We selected notes written between 1/1/2018 and 12/31/2020 and utilized the most recent note for each patient who was at least 18 years old during this period. We selected this timeframe to maximize the capture of a wide range of FDA-approved treatments. We used the most recent note per patient to avoid double counting SAEs mentioned in multiple notes, and to take advantage of the fact the documented histories tend to be inclusive of prior events. All included notes were written by a gastroenterology physician or advanced practice provider in the IBD outpatient clinic.

### Document Preprocessing

The history of present illness (HPI) section of the notes was extracted using rule-based approaches developed specifically for this project (supplemental methods). The HPI section of the note was the only portion of the note utilized for downstream analysis as this section of the note often contains a cumulative source of information on treatment exposures and outcomes, particularly out-of-system events (i.e., hospitalizations and SAEs that occurred outside of UCSF but were relayed to the gastroenterology provider at the time of routine follow-up). The HPI was pre-labeled with medications of interest, hospitalization and signs and symptoms using, named entity recognition functions, from the clinical natural language processing software *cTAKES*^12^, as well as regular expressions (i.e., the ability to locate pre-defined key-words). To minimize downstream algorithmic confusion in learning medication names, the medication brand names were replaced with the generic name using the RxNorm Application Program Interfaces (APIs) in Unified Medical Language System (UMLS)^13^.

### Note Annotation

All notes that met the above inclusion criteria were annotated to fine-tune UCSF BERT on a variety of AE detection-related tasks and to evaluate its performance against comparator models. A team of five annotators, consisting of gastroenterologists, pharmacists, pharmacovigilance experts, and patients carried out all annotation related tasks. These included the development and finalization of an annotation protocol, participation in interrater reliability assessments, and annotation of all target notes. The annotation protocol was collectively developed and refined over the course of weekly team meetings utilizing an initial subset of notes. Using LabelStudio^14^, an open-source annotation platform, annotators marked up the prelabelled HPI section of candidate notes according to triplets of medication mentions, hospitalization mentions, and SAE mentions, if they corresponded to a hospitalization as per the protocol (supplemental methods) (Figure 1). These annotations became the basis of the subsequent efforts to train UCSF BERT and other models to automate this process.

**Figure 1.**
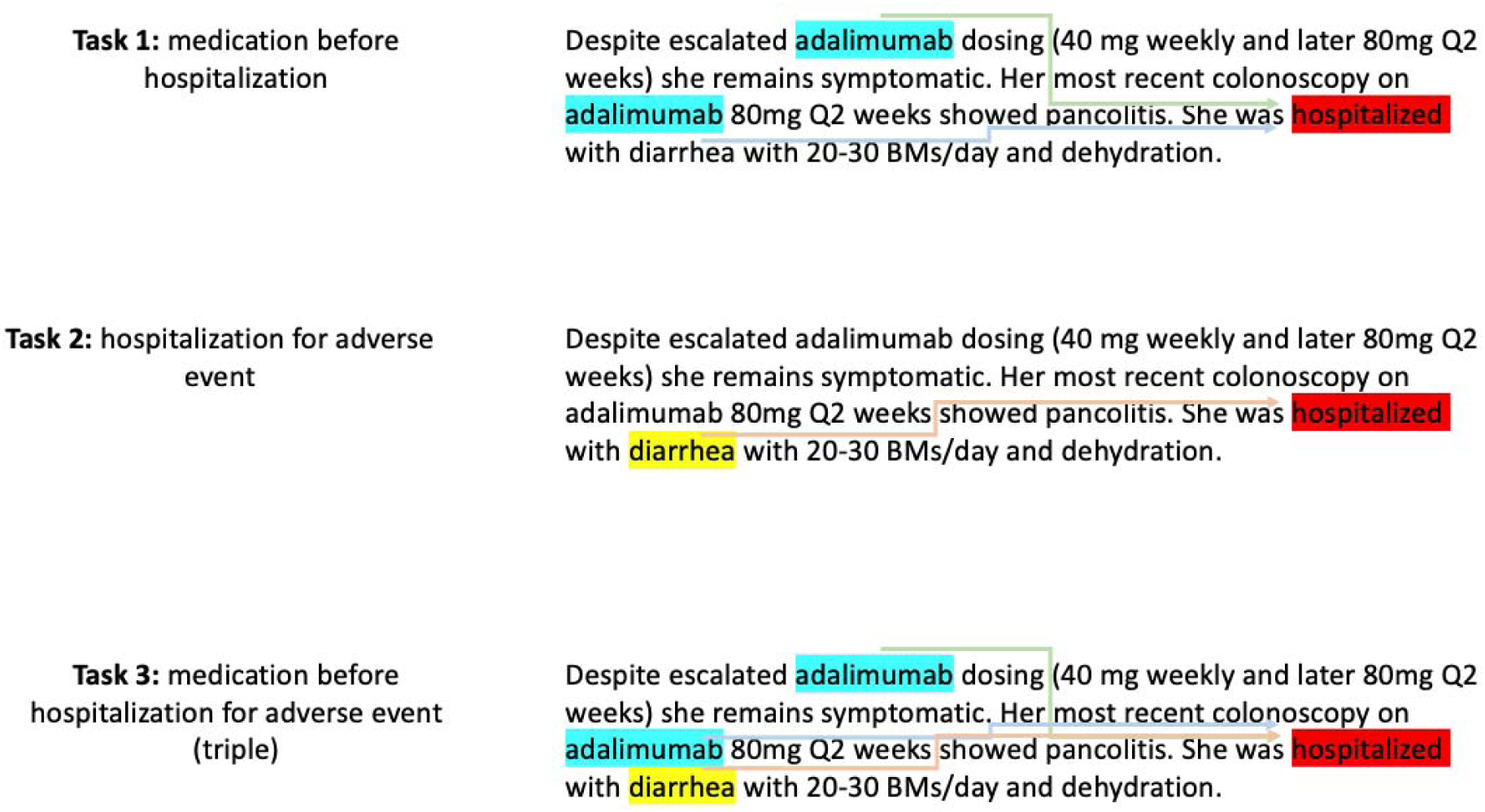
Example of the three prediction tasks from a clinical note in the corpus. Medications of interest were pre-annotated in blue, hospitalizations in red and signs and symptoms in yellow. Annotators marked up the HPI section where medications of interest predated a hospitalization (green and blue arrow) and AE causing hospitalization (red arrow). Task 3, also referred to as triple, is the combination of Task 1 and Task 2.

All five annotators participated in an interrater reliability assessment on a sample of 19 notes. The results of these assessments were reviewed in weekly meetings to improve the protocol as well as annotator compliance to it. We computed a Fleiss’ kappa statistic to characterize the interrater reliability on the final round of assessments. Following this training and assessment phase, the protocol was locked, and the remainder of the corpus was annotated.

### Modeling

We defined several prediction tasks, asking the model to classify whole HPIs according to the occurrence of: (task 1) all candidate medication mentions given prior to a hospitalization (task 2) adverse event (AE) as reason for hospitalization and (task 3) the combination of task 1 and task 2 the medication-hospitalization-AE triple (Figure 1). We trained models of different architectures to determine which were best suited for the task of AE detection. We used scikit-learn^15^ to train several baseline Bag of Words (BoW) models such as Logistic Regression, K-Nearest Neighbors, Decision Trees, Random Forest, and XGBoost (supplemental methods). We used AutoGluon^16^ to train the automated machine learning models. The annotated notes were split into 80% training, 10% validation and 10% testing. These served as a baseline to compare the performance of our UCSF BERT model. We adapted deep learning models architectures such as Convolutional Neural Network (CNN^17,18^), Bidirectional Long Short Term Memory Network (Bi-LSTM^19^) and Bi-LSTM with attention. These are deep learning models adapted from the top performing entries in the N2C2^8^ adverse event detection challenge. All BERT results are from the median performance of Macro F1 score over 5 runs of the model with different seeds. Comparative model performance significance was evaluated using Fisher’s exact test and chi square with Yates’s correlation when values were large enough to require it.

### Note Length Handling

To include the entire HPI section, which was often longer than the typical maximum input length used by other BERT models, we developed a hierarchical version of the UCSF-BERT^20^ model (H-UCSF-BERT). This model learns to process text using input sequences of 512 tokens (roughly equivalent to words), in the same manner as a typical BERT model would. It then combines them into a longer-sequence representation by integrating an additional transformer layer on top of these chunk representations. We encoded sequences up to 2560 tokens, which is 5 times the usual processing limit of a BERT model. We used the Mann-Whitney test to evaluate the possible association between note length and the presence of SAEs.

### Handling of Class Imbalance

SAEs were seen in 44% of notes in our corpus, however SAEs were uncommon once the notes were subdivided into chunks that were ingestible by H-UCSF-BERT, creating a potential problem for training models to learn to positively identify these SAEs when they do occur. To optimize learning in the face of this imbalance in the dataset, the *training data* examples without AEs were randomly undersampled. We explored a range of sampling ratios and identified the ratio of 1:4 positive to negative examples as being best for model performance. This was applied to the training dataset for all downstream tasks. We also explored additional strategies such as weighting the optimization loss based on class distributions, as well as learning these weights dynamically^21^. However, we obtained the most promising result by undersampling the majority dataset.

### MedDRA

All SAEs were manually coded using the Medical Dictionary for Regulatory Activities (MedDRA) version 23.0^22^.

## Results

### Source Corpus, Patient Population, and SAE Dataset

From a deidentified dataset of 110 million machine redacted clinical notes at UCSF, we identified a total of 928 notes corresponding to 928 adults with IBD who were seen during the 2018-2020 period. The patients in our study were 53% female with an average age of 45 years old (Table 1). The most common race of patients was white. We annotated all 928 notes and performed interrater reliability testing on a set of 19 notes to characterize the quality of the annotated dataset. The mean observed agreement among the five annotators was 93-99% across all annotation categories (Supplemental Table 1).

**Table 1.** Characteristics of patients in our note corpus.

| Patients (N=928) |  | Count(%) |
| --- | --- | --- |
| Sex | Female | 489 (52.7) |
|  | Male | 438 (47.2) |
|  | Nonbinary | 1 (0.1) |
| Age (years) | 18-40 | 460 (49.6) |
|  | 41-60 | 313 (33.7) |
|  | >60 | 155(16.7) |
| Ethnicity | Not Hispanic or Latino | 832 (89.7) |
|  | Hispanic or Latino | 83 (8.9) |
|  | Unknown/Declined | 13 (1.4) |
| Race | White or Caucasian | 670 (72.3) |
|  | Asian | 78 (8.4) |
|  | Black or African American | 41 (4.4) |
|  | American Indian or Alaska Native | 9 (1.0) |
|  | Other Pacific Islander | 1 (0.1) |
|  | Other and Unknown/Declined | 129 (13.9) |
| IBD Diagnosis | Crohn's disease | 735 (79.2) |
|  | Ulcerative colitis | 625 (67.4) |

We identified a total of 703 SAEs in the 928 annotated notes from 928 patients with IBD. All SAEs were associated with hospitalization as defined by the annotation protocol. Out of the 928 annotated notes, 411 documented at least one SAE (Table 2 and Supplemental Table 3). Importantly, some notes included more than one SAE due to multiple distinct hospitalizations in the note. The notes documenting an SAE tended to be longer than those without an SAE (p<0.001). Over 60% of SAEs in our corpus were associated with anti-tumor necrosis factor agents (anti-TNF). Infliximab was associated with 179 SAEs, the most of any drug, followed closely by adalimumab (136 SAEs; Table 3). This finding was expected given that infliximab was the first biologic to be approved for IBD, and more patients have been exposed to this medication than any other due to its longer availability. Additionally, given the relative absence of alternative treatments in the early 2000s, it is likely that patients remained on infliximab and other anti-TNFs for a longer period (even after experiencing SAEs), compared to the current era with multiple approved medications. The most common SAE was failure of intended efficacy (N=299), followed by infections (N=94) (Figure 2 and Table 4). However, SAEs were found for every organ system and every non-steroid immunosuppressant. Our corpus contained only one episode of cancer, sarcoma, which occurred in a patient receiving an anti-TNF drug. The complete list of SAEs mapped by clinical note terms and MedDRA terms can be found in the supplemental materials (Supplemental Table 3). However, to enable a more user-friendly exploration of trends in the data, we have developed an interactive web application (see https://ibd-ade.streamlit.app/).

**Figure 2.**
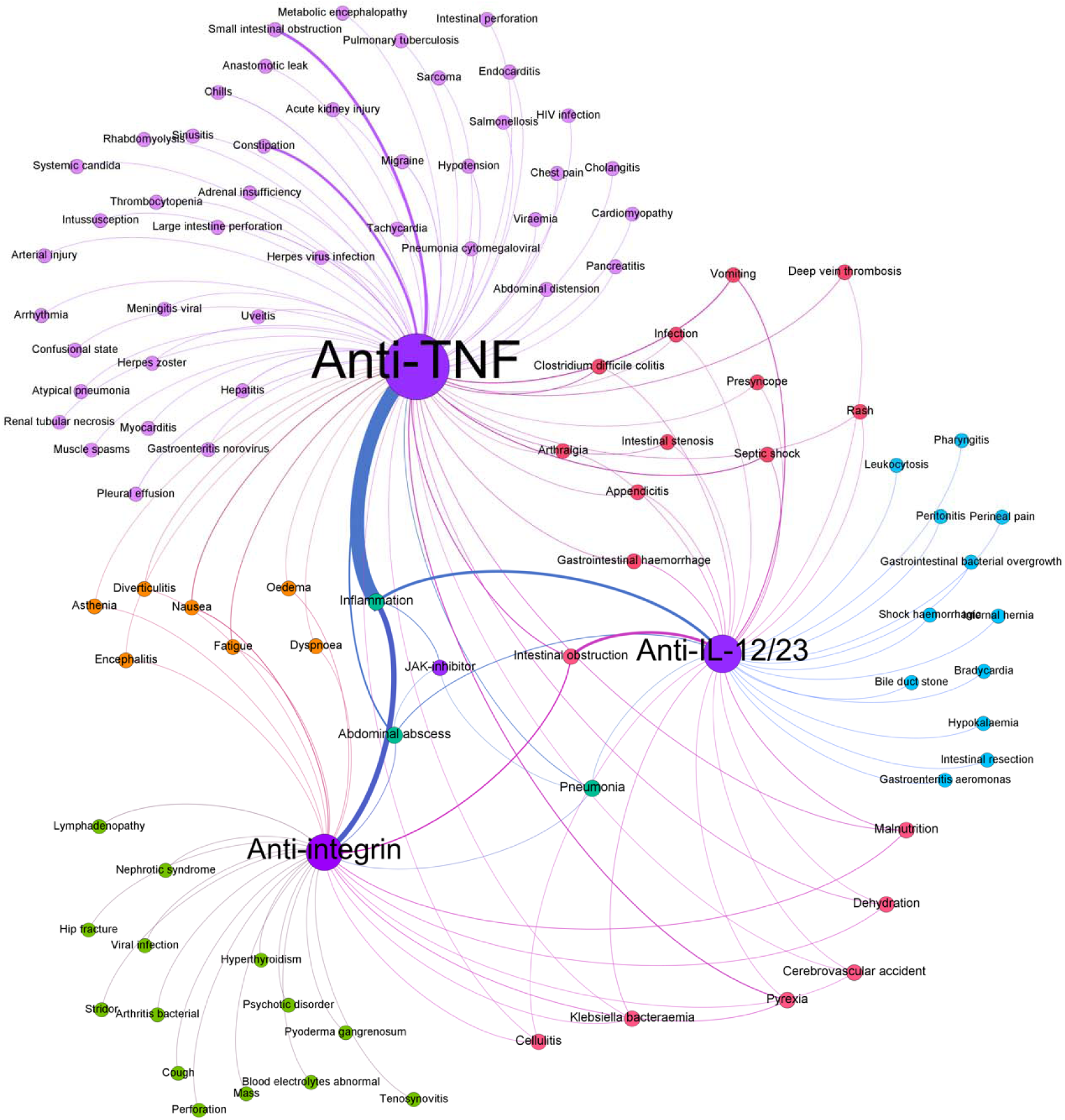
Network Graph of SAEs by medication class. The width of lines indicates the strength of association by frequency. The size of the nodes is relative to the number of exposures in our corpus to each medication. SAE colors are indicative of which medication(s) they were associated with. An interactive version of this figure can be found at https://ibd-ade.streamlit.app/

**Table 2.** Distribution of number of notes containing an SAE.

|  | Train (%) | Development (%) | Test (%) |
| --- | --- | --- | --- |
| # Annotated notes | 742 (80) | 93 (10) | 93 (10) |
| SAEs present | 335 (82) | 37 (9) | 39 (9) |
| No SAEs | 406 (79) | 56 (11) | 54 (10) |

**Table 3.** Distribution of hospitalizations and SAEs by medication. Anti-tumor necrosis factor (Anti-TNF), Janus Kinase-inhibitor (JAK-inhibitor), anti-interleukin-12/23 (anti-IL-12/23).

| Non-steroid Immunosuppressant Class | Non-steroid Immunosuppressant | Number of Hospitalizations | Number of SAEs |
| --- | --- | --- | --- |
| Anti-TNF | Adalimumab | 136 | 172 |
|  | Certolizumab | 19 | 26 |
|  | Etanercept | 2 | 1 |
|  | Golimumab | 2 | 2 |
|  | Infliximab | 179 | 231 |
| JAK-inhibitor | Tofacitinib | 8 | 8 |
| Anti-IL-12/23 | Ustekinumab | 100 | 128 |
| Anti-integrin | Vedolizumab | 106 | 135 |

**Table 4.** Top 7 SAEs in the Study. Anti-tumor necrosis factor (Anti-TNF), Janus Kinase-inhibitor (JAKi), anti-interleukin-12/23 (anti-IL-12/23).

| Serious Adverse Event MedDRA System Organ Class | Steroid Sparing Immunosuppressant Class | Frequency |
| --- | --- | --- |
| Infections and infestations | Anti-TNF | 66 |
|  | Anti-IL 12/23 | 24 |
|  | Anti-integrin | 10 |
|  | JAKi | 2 |
| Failure of intended efficacy | Anti-TNF | 216 |
|  | Anti-IL 12/23 | 75 |
|  | Anti-integrin | 90 |
|  | JAKi | 6 |
| Gastrointestinal disorders | Anti-TNF | 25 |
|  | Anti-IL 12/23 | 12 |
|  | Anti-integrin | 6 |
|  | JAKi | 0 |
| Neoplasms | Anti-TNF | 1 |
|  | Anti-IL 12/23 | 0 |
|  | Anti-integrin | 0 |
|  | JAKi | 0 |
| Cardiac disorders | Anti-TNF | 8 |
|  | Anti-IL 12/23 | 1 |
|  | Anti-integrin | 3 |
|  | JAKi | 0 |
| General disorders and administration site conditions | Anti-TNF | 21 |
|  | Anti-IL 12/23 | 1 |
|  | Anti-integrin | 9 |
|  | JAKi | 0 |
| Nervous system disorders | Anti-TNF | 7 |
|  | Anti-IL 12/23 | 2 |
|  | Anti-integrin | 2 |
|  | JAKi | 0 |

### Performance of UCSF-BERT on the Task of SAE Detection

We established three targets of prediction for all downstream models: (task 1) identify all candidate medication mentions given prior to a hospitalization, (task 2) identify adverse event as reason for hospitalization and (task 3) the combination of task 1 and task 2 the medication-hospitalization-AE triple (Figure 1). The annotated data was transformed and then split into training, validation, and testing datasets for each of these binary classification tasks (Table 2 and Supplemental Table 2). We developed and trained several variations of the UCSF-BERT model to address each of these targets. We then evaluated its performance against several other comparator models, including several of the top entries from the 2018 N2C2 adverse event detection challenge (supplemental methods).

On the task of medication prior to hospitalization, H-UCSF-BERT was the most performant model with a Macro F1 of 62% (Table 5). It was significantly more accurate than the next-best model by a margin of 11% (p < 0.01). Similarly, H-UCSF-BERT was the best model at the task of identifying hospitalization relations to AEs with an accuracy of 96% and Macro F1 of 62%. We hypothesized that long distances between mentions of a hospitalization and the associated SAEs could be reducing model accuracy. Indeed, we found that restricting the input to SAEs mentioned within a two-sentence span of the hospitalization, Macro F1 increased to 68% from 62% (p < 0.01). However, when compared to the next performant model, BiLSTM, UCSF BERT was not significantly superior (p = 0.40). The ultimate goal was to have our model accurately detect triples which include the mention of a non-steroid immunosuppressant prior to a hospitalization plus the hospitalization plus the associated SAEs. For the triples task, H-UCSF-BERT was again the best performer with a Macro F1 of 61%, however again this was not significantly different than BiLSTM (p= 0.40).

**Table 5.** Results of UCSF BERT performance on the tasks of SAE detection from real world clinical notes. Results for the three relation tasks to classify whether a pair/triple of specific entities of type medication, hospitalization and adverse event are related. Bolded models correspond to those with the best performance as measured by Macro F1. Only nearby SAEs refer to restricting only SAEs that are mentioned within a two-sentence window of the hospitalization event. Only the best three models are reported. H-UCSF-BERT = Hierarchical University of California San Francisco Bidirectional Encoder Representation from Transformers, TP = true positive, TN = true negative, FP = false positive and FN = false negative.

| Task | Model | Accuracy (%) | Macro F1 (%) | TP | TN | FP | FN |
| --- | --- | --- | --- | --- | --- | --- | --- |
| Medication before hospitalization relations | H-UCSF-BERT | 88 | 62 | 63 | 1989 | 173 | 105 |
|  | CNN | 74 | 49 | 51 | 1682 | 480 | 117 |
|  | XGBoost | 73 | 51 | 49 | 1672 | 490 | 119 |
| **Hospitalization**<br>**for SAE**<br>**relations** | H-UCSF-BERT | 96 | 62 | 79 | 9603 | 421 | 16 |
|  | **H-UCSF-BERT + only**<br>**nearby SAEs** | 92 | **68** | 34 | 1078 | 41 | 61 |
|  | BiLSTM + only nearby<br>SAEs | 93 | 48 | 7 | 1091 | 28 | 88 |
| **Medication**<br>**before**<br>**hospitalization**<br>**for SAE relation**<br>**(triples)** | **H-UCSF-BERT + only**<br>**nearby SAEs** | 91 | **61** | **141** | **7790** | **619** | **178** |
|  | CNN + only nearby AEs | 94 | 49 | 11 | 8013 | 396 | 308 |
|  | BiLSTM + only nearby<br>AEs | 95 | 50 | 11 | 7953 | 456 | 308 |

## Discussion

We adapted an EHR-specific clinical language model, UCSF BERT, to multiple tasks pertaining to the detection of treatment-emergent serious adverse events. We have evaluated its performance in the context of a specific use case: the use of non-steroid immunosuppressants for the treatment of IBD. We have generated a gold standard corpus of 928 clinical notes as the basis of training and evaluating this model against several baselines. Inter-rater reliability testing indicates good to excellent concordance across annotators. UCSF BERT performed well in a range of tasks pertaining to SAE detection from clinical notes. It achieves macro F1 scores ranging from 61-68% and accuracies from 88-92%, This model numerically outperforms existing models for SAE detection associated with the N2C2 Challenge^8^ as well as a range of strong baseline models, including several trained using automated machine learning. On the task of accurately determining a medication of interest mentioned prior to a hospitalization, UCSF BERT was significantly superior to all other models.

We found that the most common errors made by the models involved chains of reasoning across many events. For example, instances where the reason of hospitalization is not explicitly mentioned but merely implied from the clinical context. In addition, the model struggled in the setting of both long-distance dependency where there were many sentences between entities of interest and long chronology of events where several medication changes occurred over many sentences. Lastly, when there were both non-specific adverse events such as pain or vomiting as well as more specific terms such as small bowel obstruction or ulcerative colitis flare the combination was challenging for the model to handle.

The last few decades have seen a significant expansion in FDA-approved therapies for IBD. In the current era of IBD treatment with numerous agents available, continued monitoring for new safety information on these agents is helpful to inform optimal treatment selection. The most frequent SAE found in our corpus of outpatient IBD clinical notes at a tertiary referral center was failure of intended efficacy followed by infections. This is in line with previously published data, especially in the setting of more than 60% of the SAEs in our corpus associated with anti-tumor necrosis factor agents^23^. We did not account for concurrent use of steroids which are known to increase the risk of infection. However, our corpus includes SAEs from every organ system. Of note, the non-steroid medications of interest are not being prescribed with equal frequency; thus, prescribing practices are likely to influence the frequency of events as well as frequencies of possible AEs associated with the medication. The strength of association with SAEs and classes of non-steroid immunosuppressants can be explored using our interactive web application (see https://ibd-ade.streamlit.app/). The goal of developing text-based automation tools like this is to enable more precise characterizations of adverse events in the context of routine clinical care to help validate known safety profiles of these drugs as well as identify previously unrecognized SAEs which can point to areas of inquiry. For instance, our dataset included a patient receiving an anti-TNF who was hospitalized for a new diagnosis of sarcoma. Sarcomas have previously been reported in the context of children with IBD using anti-TNFs^24^, although multiple long-term observational studies have not consistently found a link between anti-TNF use and an increased risk of cancer in adults^25,26^. Future directions of this work include external validation using data from additional centers, expansion to additional disease states outside of IBD, and downstream studies designed to identify new drug-SAEs associations more rigorously using aggregated data.

Our work has many notable strengths. We have used transparent methods for developing the training corpus and assessing its quality, including interrater reliability. Because our models have been trained on de-identified clinical data, we intend to make them publicly available for others to reproduce and enhance multiple aspects of this work. Of note, the N2C2 national challenge which produced models for detection of treatment-emergent adverse events from clinical notes prior to our work was before the release of BERT. We suspect that the underlying architecture of BERT in addition to our pre-training from scratch on a sizeable clinical corpus are driving our improved performance compared to prior models.

Some limitations of our work, outside of those common to retrospective research, include imperfect accuracy of the model, which on certain tasks did not perform statistically significantly superior to other models. We suspect this largely the result of long-distance dependance and long chains of reasoning across many events in a clinical note. In addition, we have not yet assessed the generalizability of our model across other diseases, treatments, or health systems. As well, our interrater agreement is potentially optimistic as it was calculated iteratively on the same 19 notes. However, there are no universally accepted standards of the Fleiss’ kappa statistic^27^ for good agreement. Future work aimed at improving upon our current model includes annotating a lager corpus at an outside health system to evaluate generalizability and over-sampling for SAEs to have more positive examples for the model to learn from. Overall, our approach, utilizing novel methods from the field of artificial intelligence, has the potential to address unmet needs in drug safety surveillance, an area of central importance to regulatory agencies across the globe and to public health in general.

## Conclusion

We have successfully adapted a new clinical language model, UCSF BERT, to the task of mining outpatient clinic notes for SAEs occurring in patients with IBD administered non-steroid immunosuppressants. This model performs well on the tasks of SAE detection, especially identifying target medications prior to hospitalizations. The success of this model appears to stem from its pretraining on a large and diverse corpus of notes derived from real-world clinical care and use of hierarchical modeling which allows for long sequence document classification tasks. These results suggest the feasibility of adapting artificial intelligence methods to address important unmet needs in the field of pharmacovigilance, with the potential to substantially reduce the manual efforts needed to review notes and identify events of concern. Our work is a step closer to a future of automated drug surveillance algorithms embedded within EHR systems which can facilitate pharmacovigilance activities ranging from health system reporting of SAEs to large-scale safety evaluations across multiple EHR systems without the limitations of using billing codes as surrogates for actual AEs.

## Study Highlights

- What is the current knowledge on the topic?

- Prior work in automated adverse event (AE) detection from routine clinic notes utilized note fragments and simplified AE detection tasks. In addition, the newest model architectures were not widely available when automated AE detection was evaluated in a national natural language processing challenge in 2018.
- What question did this study address?

- Are the newest model architectures trained on clinical notes capable of detecting serious AEs in routine clinical notes as written by clinicians seeing patients with inflammatory bowel diseases at a tertiary medical center.
- What does this study add to our knowledge?

- Our model, UCSF BERT, trained on a large corpus of real-world clinical notes performs better than prior models previously designed for this task. Notably, our hierarchical model architecture is able to digest information five times the usual processing limit of BERT.
- How might this change clinical pharmacology or translational science?

- Our work is a step closer to a future of automated drug surveillance algorithms embedded within EHR systems which can facilitate pharmacovigilance activities.

## Access to Data

The analytic code to train and evaluate models will be made publicly available at https://github.com/MadhumitaSushil/ADE_detection. A machine-redacted version of the notes-based data can be made available to requesting researchers by mutual agreement and following the execution of a data use agreement.

## Acknowledgements

We gratefully acknowledge the invaluable administrative support provided by Lily Wong. In addition, we would like to acknowledge the <u>UCSF Information Commons Computational Research</u> <u>Platform</u>, developed and supported by UCSF Bakar Computational Health Sciences Institute and UCSF Academic Research Services. We would like to express our thanks to the Wynton support team and the UCSF high-performance computing cluster, Wynton.

## Involvement with the Manuscript

Anna L. Silverman: study concept and design; acquisition of data; analysis and interpretation of data; lead annotation protocol; lead drafting of the manuscript; lead critical revision of the manuscript for important intellectual content

Madhumita Sushil: co-lead model architect, study concept and design; acquisition of data; analysis and interpretation of data; drafting of the manuscript; critical revision of the manuscript for important intellectual content; technical support; implementation and model design

Balu Bhasuran: co-lead model architect, study concept and design; acquisition of data; analysis and interpretation of data; drafting of the manuscript; critical revision of the manuscript for important intellectual content; technical support; implementation and model design

Dana Ludwig: study concept and design; acquisition of data; drafting of the manuscript; critical revision of the manuscript for important intellectual content; technical support; implementation and model design

James Buchanan: study concept and design; acquisition of data; analysis and interpretation of data; critical revision of the manuscript for important intellectual content

Rebecca Racz: study concept and design; acquisition of data; analysis and interpretation of data; critical revision of the manuscript for important intellectual content

Mahalakshmi Parakala: acquisition of data

Samer El-Kamary: lead investigator at the FDA for this project, administrative responsibility and study team supervision at the FDA, intellectual contribution during the conduct of the study, and critical revision of the manuscript for important intellectual content.

Ohenewaa Ahima: critical revision of the manuscript for important intellectual content; study supervision

Artur Belov: critical revision of the manuscript for important intellectual content; study supervision

Lauren Choi: critical revision of the manuscript for important intellectual content; study supervision

Monisha Billings: critical revision of the manuscript for important intellectual content; study supervision

Yan Li: critical revision of the manuscript for important intellectual content; study supervision

Nadia Habal: critical revision of the manuscript for important intellectual content; study supervision

Qi Liu: critical revision of the manuscript for important intellectual content; study supervision

Jawahar Tiwari: critical revision of the manuscript for important intellectual content; study supervision

Atul Butte: study supervision

Vivek A Rudrapatna: study concept and design; acquisition of data; analysis and interpretation of data; technical support; critical revision of the manuscript for important intellectual content; study supervision

## Disclaimer

The contents of this article reflect the views of the authors and should not be construed to represent the FDA’s views or policies. No official support or endorsement by the FDA is intended or should be inferred.

