## Supplemental material for "Algorithmic identification of treatment-emergent adverse events from clinical notes using large language models: a pilot study in inflammatory bowel disease"

**Supplemental Materials**

### Non-steroid Immunosuppressant Target Medications

- Infliximab (Remicade) approved by the FDA August 1998
- Infliximab-dyyb (Inflectra or Remsima)
- Adalimumab (Humira) approved by the FDA January 2008
- Golimumab (Simponi) approved by the FDA May 2013
- Certolizumab Pegol (Cimzia) approved by the FDA April 2008
- Natalizumab (Tysabri) approved by the FDA November 2004
- Vedolizumab (Entyvio) approved by the FDA May 2014
- Ustekinumab (Stelara) approved by the FDA September 2016
- Tofacitinib (Xeljanz) approved by the FDA May 2018
- Upadacitinib (Rinvoq) approved by the FDA March 2022
- Etanercept (Enbrel) approved by the FDA July 2003

### Limiting Notes to the History of Present Illness

- The beginning of the HPI was defined by the regex pattern:
  - r'(.*?)(History of Present Illness|HPI)(.*$)'
  - if the pattern was not found, we included text from the beginning of the progress note
- The end of the HPI was defined by the regex pattern:
  - r"(.*?)(Past Medical History:|PMH:|PMHx:|Medical/Surgical History|" \

+ r"Review of Systems|Current Outpatient Medications on File|" \

+ r"Assessment and Plan|\bROS\b)(.*$)"

- If the pattern was not found, we included the text to the end of the progress note.
- In the HPI text, all sequences of more than 1 blank space were replaced with one blanks space

### Pre-annotating all Mentions of Steroid Sparing Immunosuppressants and Hospitalizations

- The medication mentions were those that matched the name of either a trade name or generic name of one of the biologics in the table above. For each med string mention, the generic name was recorded from the table above.
- The hospitalization mentions were found using the regex pattern:
  - r'(admitted|admited|hospital|admission)'

### Annotation Protocol

- Annotators were asked to complete three tasks per note:
  - Task 1: Find a hospitalization that occurred AFTER starting a target medication. Link all mentions of the same hospitalization together.
  - Task 2: Associate that hospitalization with every mention of the target medication used prior to hospital admission
  - Task 3: Associate the hospitalization with the reason for hospitalization
- If ambiguity was encountered annotators kept a list of notes to discuss as a group and adjudicate on a case-by-case basis. Adjudication meetings were held weekly.

###### 1. Annotation Protocol

Task 1: Find hospitalizations that occur after starting medications of interest but before stopping the medications of interest. Link all mentions of the same hospitalization together.

1. Look at every hospitalization to determine if the hospitalization occurred AFTER starting a medication of interest
2. If a medication of interest was NOT started prior to the first hospitalization then move on to the next hospitalization until you find a hospitalization that occurs AFTER starting a medication of interest or all hospitalizations in the note have been reviewed and none occurred in the setting of medications of interest use.
3. The hospitalization must occur AFTER starting a medication of interest but BEFORE the decision is made to stop the medication of interest
4. If there are two or more words indicating the same hospitalization in the same or different sentences, they should be linked together in a path-connected fashion by relationship arrows.

Once a hospitalization is found that occurred after the start of a medication of interest but before stopping the medication of interest and all assertions of this same hospitalization have been linked to each other proceed to Task 2.

There may be multiple hospitalizations in a single note that occur after the start of a medication of interest. EVERY hospitalization that occurs AFTER a biologic has started but before stopping the biologic should be annotated.

If a hospitalization is found that is not pre-annotated in red this can be done manually by clicking
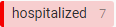
 and then using your cursor to highlight the word or phrase indicating hospitalization occurred.

Task 2: Link hospitalizations that occur after starting a medication of interest with the specific medication of interest used

1. Click on medication of interest name that is the medication of interest at time of hospitalization. If two medications of interest are being used concurrently prior to the hospitalization (this is rare) BOTH should be linked (this requires repeating this step individually for each biologic).
2. Click
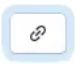
 icon on right hand side of screen. Also, can use short cut by holding down the “r” key on keyboard.
3. Click on hospitalization. A red bracket should appear between the medication of interest and hospitalization indicating the correct relationship has been made.
   a. The direction of the red arrow does not matter
   b. If the medication of interest is mentioned in the opening summary line of the note it should NOT be linked to the hospitalization.
4. Repeat this process for ALL mentions of the medication(s) of interest and link to the hospitalization(s).
   a. If there are multiple mentions of the same hospitalization, as long as all mentions of the same hospitalization are linked in a path connected fashion from Task 1 then all mentions of treatment-emergent medication(s) of interest need only be linked to one mention of the hospitalization.
   b. If there are multiple treatment associated hospitalizations this process should be repeated for each.

Task 3: Link the hospitalization that occurred after biologic started with the reason for hospitalization. If no reason for hospitalization then DO NOT LABEL ANYTHING!

1. Click on the reason for hospitalization. This should be the highest-level diagnosis(s) that led to hospitalization and must occur within 2 sentences before or after the mention of the hospitalization. If the reason for hospitalization is mentioned multiple times within 2 sentences before or after hospitalization then the synonymous reasons for hospitalization should be connected to each other.
   a. If available, prefer more specific diagnosis (Ex. Prefer small bowel obstruction rather than obstruction. Ex prefer salmonella infections rather than infection.)
   b. If IBD (ulcerative colitis, Crohn’s, colitis) is the reason for hospitalization then pick this. However, if “flare” is all that is used then you can highlight flare. Otherwise DO NOT include flare in the annotation.
   c. DO NOT include adjectives such as severe or acute in the annotation of reason for hospitalization.
   d. If no diagnosis is indicated then it is acceptable to label symptom(s) as a reason(s) for hospitalization.
   e. If no diagnosis or symptoms are indicated as a reason for hospitalization then DO NOT annotate anything.
   f. Treatments or test results, such as surgeries, should not be labeled as reasons for hospitalization unless a diagnosis is stated from them ie. C dif positive or CT consistent with flare of colitis
   g. If the reason for hospitalization is redacted, DO NOT label it as the reason for hospitalization.
2. Click
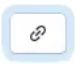
 icon on right hand side of screen. Also, can use short cut with “r” key on keyboard.
3. Click on hospitalization. A red bracket should appear between the reason for hospitalization and hospitalization indicating the correct relationship has been made.
   a. The direction of the red arrow does not matter

### Modeling

- We used machine learning models using Bag-of-Words (BoW) method and chosen high performance models such as Logistic Regression, K-Nearest Neighbors, Decision Tree, Random Forest and XGBoost. These served as a baseline to compare the performance of our UCSF BERT.
- We adapted deep learning models architectures such as, Convolutional Neural Network (CNN^17,18^), Bidirectional Long Short Term Memory Network (Bi-LSTM^19^) and Bi-LSTM + Attention. These are deep learning models adapted from prior N2C2 tasks and prior to our UCSF BERT model held the state-of-the art status for the task of adverse event detection.
- The baseline model parameters were set to default values and then optimized using grid search. For the deep learning models, hyperparameters were optimized using an incremental approach (e.g. learning rate 0.1, 0.01, 0.001). Hyperparameters in the autoML-trained models were algorithmically optimized.
- The models were generated in Python 3.8 using Scikit-learn 1.1^15^ for baseline models, deep learning models were generated using PyTorch 1.11.0^28^, and autoML models were generated using AutoGluon 0.4.2^16^ packages respectively.

### Inference Task Set Up

- As the first task, we have implemented a classifier that takes the HPI section of a note as the input, and outputs whether it contains the mention of a SAE, which was determined as discussed earlier.
- In the next set of tasks, we have implemented relation extraction classifiers to determine relations between pairs of specific events. Mentions of the individual events of interest are replaced with placeholders, and the model is asked to infer whether a valid relation exists between these placeholders. In the first relation classification task, we aim to find whether the patient was on the medication @MED$ before the hospitalization @HOSP$ happened. Here, @MED$ is the placeholder for the medication of interest, and @HOSP$ is the placeholder for a specific hospitalization event.
- In the second relation classification task, we have trained the model to classify whether a specific adverse event mentioned in the text, represented by the @AE$ token, was the cause of a hospitalization event denoted @HOSP$. These tasks are based on the relations that were added by the annotators while curating the annotated dataset. We hypothesize that we can obtain a better performance by disintegrating the task of SAE detection into these sub-tasks, as we alleviate the need for any causal inferences at this step.
- In the final task, we ask the model to classify relations between triples of events: a target medication mention, a hospitalization mention, and an adverse event. The relation is present if both the target medication and the adverse event are related to the same hospitalization event, absent otherwise. We use the UCSF-BERT model for performing classification as well as for inferring relations described above.

### Clinical Terms and Corresponding MedDRA Codes of SAEs in Our Corpus

- When the Clinical Term(s) containing more than one medical concept, each is separately coded to an individual MedDRA Preferred Term (PT).
- All coding using MedDRA version 23.0

### Inter-annotator Agreement

**Supplemental Table 1.** Inter-rater reliability assessment results

| **Annotation** | **Type** | **Observed Agreement** | **Chance Agreement** | **Fleiss’ Kappa** |
| --- | --- | --- | --- | --- |
| Concept | AE | 99% | 96% | 0.62 |
| Concept | Hospitalized | 93% | 53% | 0.84 |
| Concept | Med | 95% | 78% | 0.77 |
| Relations | Med before hosp | 97% | 87% | 0.76 |
| Relations | Hosp for AE | 99% | 98% | 0.62 |

### Number of Positive and Negative Relations for Different Relation Classification Subtasks

**Supplemental Table 2.** Number of positive and negative relations for different relation classification subtask triplets (medication of interest, hospitalization, and AE) in the source corpus

|  | **Med before hospitalization** | | | **Hosp for AE** | | | **Serious AE relations** | | |
| --- | --- | --- | --- | --- | --- | --- | --- | --- | --- |
|  | Train (%) | Development (%) | Test (%) | Train (%) | Dev (%) | Test (%) | Train (%) | Dev (%) | Test (%) |
| **Positive relations** | 1396 (82) | 128 (8) | 168 (10) | 858 (84) | 69 (7) | 95 (9) | 1886 (78) | 209 (9) | 319 (13) |
| **Negative relations** | 15377 (80) | 1814 (9) | 2162 (11) | 9594 (81) | 1202 (10) | 1119 (9) | 57043 (80) | 6322 (8) | 8409 (12) |

### SAE Frequencies and Mapping by Clinical Note Terms, MedDRA Terms and Medication

**Supplemental Table 3.** SAE frequencies mapped by clinical note terms, MedDRA preferred terms and medication

| **Serious Adverse Event MedDRA System Organ Class** | **Note Clinical Term(s)** | **MedDRA Preferred Term(s)** | **Medication** | **Frequency** |
| --- | --- | --- | --- | --- |
| **Infections and infestations** | C diff = c. difficile colitis = c. difficile = infectious colitis | Clostridium difficile colitis | Infliximab | 5 |
|  |  | Enterocolitis infectious | Adalimumab | 1 |
|  |  |  | Certolizumab | 0 |
|  |  |  | Golimumab | 0 |
|  |  |  | Etanercept | 0 |
|  |  |  | Ustekinumab | 2 |
|  |  |  | Vedolizumab | 0 |
|  |  |  | Tofacitnib | 0 |
|  | Campylobacter | Campylobacter infection | Infliximab | 0 |
|  |  |  | Adalimumab | 0 |
|  |  |  | Certolizumab | 0 |
|  |  |  | Golimumab | 0 |
|  |  |  | Etanercept | 0 |
|  |  |  | Ustekinumab | 1 |
|  |  |  | Vedolizumab | 0 |
|  |  |  | Tofacitnib | 0 |
|  | Pneumonia = CAP (community acquired pneumonia) = PNA | Pneumonia | Infliximab | 4 |
|  |  |  | Adalimumab | 1 |
|  |  |  | Certolizumab | 0 |
|  |  |  | Golimumab | 0 |
|  |  |  | Etanercept | 0 |
|  |  |  | Ustekinumab | 1 |
|  |  |  | Vedolizumab | 1 |
|  |  |  | Tofacitnib | 1 |
|  | Gram negative rod bacteremia = Enterococcus bacteremia = Klebsiella bacteremia = bacteremia = staph epi bacteremia | Bacteraemia | Infliximab | 0 |
|  |  | Klebsiella bacteraemia | Adalimumab | 0 |
|  |  | Staphylococcal bacteraemia | Certolizumab | 1 |
|  |  |  | Golimumab | 0 |
|  |  |  | Etanercept | 0 |
|  |  |  | Ustekinumab | 2 |
|  |  |  | Vedolizumab | 1 |
|  |  |  | Tofacitnib | 0 |
|  | Abscess = Abdominal abscess = pelvic abscess = liver abscess = perirectal abscess = gluteal abscess = abscesses = perianal abscess = intra-abdominal abcess | Abscess | Infliximab | 8 |
|  |  | Abdominal abscess | Adalimumab | 11 |
|  |  | Pelvic abscess | Certolizumab | 0 |
|  |  | Liver abscess | Golimumab | 0 |
|  |  | Perirectal abscess | Etanercept | 0 |
|  |  | Abscess limb | Ustekinumab | 7 |
|  |  | Anal abscess | Vedolizumab | 4 |
|  |  | Abdominal abscess | Tofacitnib | 1 |
|  | UTI = urinary tract infection = pyelonephritis | Urinary tract infection | Infliximab | 1 |
|  |  | Pyelonephritis | Adalimumab | 1 |
|  |  |  | Certolizumab | 0 |
|  |  |  | Golimumab | 0 |
|  |  |  | Etanercept | 0 |
|  |  |  | Ustekinumab | 1 |
|  |  |  | Vedolizumab | 0 |
|  |  |  | Tofacitnib | 0 |
|  | septic = septic shock = sepsis = urosepsis | Septic shock | Infliximab | 4 |
|  |  | Sepsis | Adalimumab | 4 |
|  |  | Urosepsis | Certolizumab | 1 |
|  |  |  | Golimumab | 0 |
|  |  |  | Etanercept | 0 |
|  |  |  | Ustekinumab | 3 |
|  |  |  | Vedolizumab | 0 |
|  |  |  | Tofacitnib | 0 |
|  | septic arthritis | Arthritis bacterial | Infliximab | 0 |
|  |  |  | Adalimumab | 0 |
|  |  |  | Certolizumab | 0 |
|  |  |  | Golimumab | 0 |
|  |  |  | Etanercept | 0 |
|  |  |  | Ustekinumab | 0 |
|  |  |  | Vedolizumab | 1 |
|  |  |  | Tofacitnib | 0 |
|  | pulmonary tuberculosis | Pulmonary tuberculosis | Infliximab | 1 |
|  |  |  | Adalimumab | 0 |
|  |  |  | Certolizumab | 0 |
|  |  |  | Golimumab | 0 |
|  |  |  | Etanercept | 0 |
|  |  |  | Ustekinumab | 0 |
|  |  |  | Vedolizumab | 0 |
|  |  |  | Tofacitnib | 0 |
|  | endocarditis | Endocarditis | Infliximab | 1 |
|  |  |  | Adalimumab | 0 |
|  |  |  | Certolizumab | 0 |
|  |  |  | Golimumab | 0 |
|  |  |  | Etanercept | 0 |
|  |  |  | Ustekinumab | 0 |
|  |  |  | Vedolizumab | 0 |
|  |  |  | Tofacitnib | 0 |
|  | staph infection | Staphylococcal infection | Infliximab | 0 |
|  |  |  | Adalimumab | 1 |
|  |  |  | Certolizumab | 0 |
|  |  |  | Golimumab | 0 |
|  |  |  | Etanercept | 0 |
|  |  |  | Ustekinumab | 0 |
|  |  |  | Vedolizumab | 0 |
|  |  |  | Tofacitnib | 0 |
|  | sinus infection | Sinusitis | Infliximab | 1 |
|  |  |  | Adalimumab | 0 |
|  |  |  | Certolizumab | 0 |
|  |  |  | Golimumab | 0 |
|  |  |  | Etanercept | 0 |
|  |  |  | Ustekinumab | 0 |
|  |  |  | Vedolizumab | 0 |
|  |  |  | Tofacitnib | 0 |
|  | Cellulitis | Cellulitis | Infliximab | 1 |
|  |  |  | Adalimumab | 0 |
|  |  |  | Certolizumab | 0 |
|  |  |  | Golimumab | 0 |
|  |  |  | Etanercept | 0 |
|  |  |  | Ustekinumab | 1 |
|  |  |  | Vedolizumab | 1 |
|  |  |  | Tofacitnib | 0 |
|  | aeromonas gastroenteritis | Gastroenteritis aeromonas | Infliximab | 0 |
|  |  |  | Adalimumab | 0 |
|  |  |  | Certolizumab | 0 |
|  |  |  | Golimumab | 0 |
|  |  |  | Etanercept | 0 |
|  |  |  | Ustekinumab | 1 |
|  |  |  | Vedolizumab | 0 |
|  |  |  | Tofacitnib | 0 |
|  | pharyngitis | Pharyngitis | Infliximab | 1 |
|  |  |  | Adalimumab | 0 |
|  |  |  | Certolizumab | 0 |
|  |  |  | Golimumab | 0 |
|  |  |  | Etanercept | 0 |
|  |  |  | Ustekinumab | 0 |
|  |  |  | Vedolizumab | 0 |
|  |  |  | Tofacitnib | 0 |
|  | line infection | Vascular device infection | Infliximab | 0 |
|  |  |  | Adalimumab | 0 |
|  |  |  | Certolizumab | 0 |
|  |  |  | Golimumab | 0 |
|  |  |  | Etanercept | 0 |
|  |  |  | Ustekinumab | 1 |
|  |  |  | Vedolizumab | 0 |
|  |  |  | Tofacitnib | 0 |
|  | salmonella | Salmonellosis | Infliximab | 0 |
|  |  |  | Adalimumab | 1 |
|  |  |  | Certolizumab | 0 |
|  |  |  | Golimumab | 0 |
|  |  |  | Etanercept | 0 |
|  |  |  | Ustekinumab | 0 |
|  |  |  | Vedolizumab | 0 |
|  |  |  | Tofacitnib | 0 |
|  | atypical pneumonia | Atypical pneumonia | Infliximab | 1 |
|  |  |  | Adalimumab | 0 |
|  |  |  | Certolizumab | 0 |
|  |  |  | Golimumab | 0 |
|  |  |  | Etanercept | 0 |
|  |  |  | Ustekinumab | 0 |
|  |  |  | Vedolizumab | 0 |
|  |  |  | Tofacitnib | 0 |
|  | Zoster | Herpes zoster | Infliximab | 0 |
|  |  |  | Adalimumab | 1 |
|  |  |  | Certolizumab | 0 |
|  |  |  | Golimumab | 0 |
|  |  |  | Etanercept | 0 |
|  |  |  | Ustekinumab | 0 |
|  |  |  | Vedolizumab | 0 |
|  |  |  | Tofacitnib | 0 |
|  | HSV | Herpes virus infection | Infliximab | 2 |
|  |  |  | Adalimumab | 0 |
|  |  |  | Certolizumab | 0 |
|  |  |  | Golimumab | 0 |
|  |  |  | Etanercept | 0 |
|  |  |  | Ustekinumab | 0 |
|  |  |  | Vedolizumab | 0 |
|  |  |  | Tofacitnib | 0 |
|  | CMV pneumonitis = CMV viremia = CMV | Pneumonia cytomegaloviral | Infliximab | 1 |
|  |  | Cytomegalovirus viraemia | Adalimumab | 1 |
|  |  | Cytomegalovirus infection | Certolizumab | 1 |
|  |  |  | Golimumab | 0 |
|  |  |  | Etanercept | 0 |
|  |  |  | Ustekinumab | 0 |
|  |  |  | Vedolizumab | 0 |
|  |  |  | Tofacitnib | 0 |
|  | norovirus | Gastroenteritis norovirus | Infliximab | 1 |
|  |  |  | Adalimumab | 0 |
|  |  |  | Certolizumab | 0 |
|  |  |  | Golimumab | 0 |
|  |  |  | Etanercept | 0 |
|  |  |  | Ustekinumab | 0 |
|  |  |  | Vedolizumab | 0 |
|  |  |  | Tofacitnib | 0 |
|  | viral | Viral infection | Infliximab | 0 |
|  |  |  | Adalimumab | 0 |
|  |  |  | Certolizumab | 0 |
|  |  |  | Golimumab | 0 |
|  |  |  | Etanercept | 0 |
|  |  |  | Ustekinumab | 0 |
|  |  |  | Vedolizumab | 1 |
|  |  |  | Tofacitnib | 0 |
|  | HIV | HIV infection | Infliximab | 1 |
|  |  |  | Adalimumab | 0 |
|  |  |  | Certolizumab | 0 |
|  |  |  | Golimumab | 0 |
|  |  |  | Etanercept | 0 |
|  |  |  | Ustekinumab | 0 |
|  |  |  | Vedolizumab | 0 |
|  |  |  | Tofacitnib | 0 |
|  | viral meningitis | Meningitis viral | Infliximab | 1 |
|  |  |  | Adalimumab | 0 |
|  |  |  | Certolizumab | 0 |
|  |  |  | Golimumab | 0 |
|  |  |  | Etanercept | 0 |
|  |  |  | Ustekinumab | 0 |
|  |  |  | Vedolizumab | 0 |
|  |  |  | Tofacitnib | 0 |
|  | viremia | Viraemia | Infliximab | 1 |
|  |  |  | Adalimumab | 0 |
|  |  |  | Certolizumab | 0 |
|  |  |  | Golimumab | 0 |
|  |  |  | Etanercept | 0 |
|  |  |  | Ustekinumab | 0 |
|  |  |  | Vedolizumab | 0 |
|  |  |  | Tofacitnib | 0 |
|  | infection | Infection | Infliximab | 1 |
|  |  |  | Adalimumab | 0 |
|  |  |  | Certolizumab | 0 |
|  |  |  | Golimumab | 0 |
|  |  |  | Etanercept | 0 |
|  |  |  | Ustekinumab | 1 |
|  |  |  | Vedolizumab | 0 |
|  |  |  | Tofacitnib | 0 |
|  | candidemia | Systemic candida | Infliximab | 1 |
|  |  |  | Adalimumab | 0 |
|  |  |  | Certolizumab | 0 |
|  |  |  | Golimumab | 0 |
|  |  |  | Etanercept | 0 |
|  |  |  | Ustekinumab | 0 |
|  |  |  | Vedolizumab | 0 |
|  |  |  | Tofacitnib | 0 |
|  | appendicitis | Appendicitis | Infliximab | 1 |
|  |  |  | Adalimumab | 1 |
|  |  |  | Certolizumab | 0 |
|  |  |  | Golimumab | 0 |
|  |  |  | Etanercept | 0 |
|  |  |  | Ustekinumab | 2 |
|  |  |  | Vedolizumab | 0 |
|  |  |  | Tofacitnib | 0 |
|  | sigmoid diverticulitis = diverticulitis | Diverticulitis | Infliximab | 2 |
|  |  |  | Adalimumab | 0 |
|  |  |  | Certolizumab | 0 |
|  |  |  | Golimumab | 0 |
|  |  |  | Etanercept | 0 |
|  |  |  | Ustekinumab | 0 |
|  |  |  | Vedolizumab | 1 |
|  |  |  | Tofacitnib | 0 |
|  | small intestinal bacterial overgrowth = SIBO | Gastrointestinal bacterial overgrowth | Infliximab | 0 |
|  |  |  | Adalimumab | 0 |
|  |  |  | Certolizumab | 0 |
|  |  |  | Golimumab | 0 |
|  |  |  | Etanercept | 0 |
|  |  |  | Ustekinumab | 1 |
|  |  |  | Vedolizumab | 0 |
|  |  |  | Tofacitnib | 0 |
| ***Failure of Intended Efficacy*** | inflammation = flare = flares = flaring = flared = proctitis = ulcerative colitis = severe abdominal pain = abdominal pain = diarrhea = loose stools = 20 BM per day = bleeding = Iron deficiency = anemia = disease = maroon stools = hematochezia = colitis = disease activity = refractory disease = frequent stools = >15 bloody bowel movements = RLQ pain = right lower quadrant pain = RUQ abdominal pain = epigastric pain = LLQ pain = LLQ abdominal pain = left lower abdominal pain = rectal pain = tenesmus = abdominal crampy abdominal pain = inflammatory bowel disease = blood in her stool = ileitis = acute ulcerative colitis = severe lower abdominal pain = blood stool = bloody stool = crohns = crohn = pelvic pain = acute abdominal pain = severe inflammation = bloody diarrhea = stool frequency &#10 increased = cramping = enteritis = IBD = ulceration = ulcer = pancolitis = increased stool frequency = colon ulceration = active inflammation = acute inflammation = rectal bleeding = bowel movements = UC flare = urgency | Inflammation | Infliximab | 102 |
|  |  | Proctitis | Adalimumab | 74 |
|  |  | Colitis ulcerative | Certolizumab | 9 |
|  |  | Abdominal pain | Golimumab | 2 |
|  |  | Diarrhoea | Etanercept | 1 |
|  |  | Haemorrhage | Ustekinumab | 35 |
|  |  | Iron deficiency anaemia | Vedolizumab | 70 |
|  |  | Haematochezia | Tofacitnib | 6 |
|  |  | Colitis |  |  |
|  |  | Abdominal pain lower |  |  |
|  |  | Abdominal pain upper |  |  |
|  |  | Proctalgia |  |  |
|  |  | Rectal tenesmus |  |  |
|  |  | Inflammatory bowel disease |  |  |
|  |  | Crohn's disease |  |  |
|  |  | Pelvic pain |  |  |
|  |  | Diarrhoea haemorrhagic |  |  |
|  |  | Frequent bowel movements |  |  |
|  |  | Muscle spasms |  |  |
|  |  | Enteritis |  |  |
|  |  | Ulcer |  |  |
|  |  | Colitis |  |  |
|  |  | Rectal haemorrhage |  |  |
|  |  | Colitis ulcerative |  |  |
|  |  | Defaecation urgency |  |  |
|  | small bowel obstruction = SBO = obstruction = obstipation = obstructive symptoms = bowel obstructions = bowel obstruction = obstructions = partial bowel obstruction = small bowel &#10 obstruction | Small intestinal obstruction | Infliximab | 36 |
|  |  | Constipation | Adalimumab | 31 |
|  |  | Intestinal obstruction | Certolizumab | 9 |
|  |  |  | Golimumab | 0 |
|  |  |  | Etanercept | 0 |
|  |  |  | Ustekinumab | 34 |
|  |  |  | Vedolizumab | 13 |
|  |  |  | Tofacitnib | 0 |
|  | fistula = enteroenteric fistula = enterocutaneous fistula = perianal disease = sinus tract = active fistulizing disease | Fistula | Infliximab | 0 |
|  |  | Enterocutaneous fistula | Adalimumab | 1 |
|  |  |  | Certolizumab | 0 |
|  |  |  | Golimumab | 0 |
|  |  |  | Etanercept | 0 |
|  |  |  | Ustekinumab | 2 |
|  |  |  | Vedolizumab | 6 |
|  |  |  | Tofacitnib | 0 |
|  | stricture | Intestinal stenosis | Infliximab | 1 |
|  |  |  | Adalimumab | 2 |
|  |  |  | Certolizumab | 0 |
|  |  |  | Golimumab | 0 |
|  |  |  | Etanercept | 0 |
|  |  |  | Ustekinumab | 1 |
|  |  |  | Vedolizumab | 0 |
|  |  |  | Tofacitnib | 0 |
|  | perineal pain | Perineal pain | Infliximab | 0 |
|  |  |  | Adalimumab | 0 |
|  |  |  | Certolizumab | 0 |
|  |  |  | Golimumab | 0 |
|  |  |  | Etanercept | 0 |
|  |  |  | Ustekinumab | 1 |
|  |  |  | Vedolizumab | 0 |
|  |  |  | Tofacitnib | 0 |
|  | ileocolonic resection | Intestinal resection | Infliximab | 0 |
|  |  |  | Adalimumab | 0 |
|  |  |  | Certolizumab | 0 |
|  |  |  | Golimumab | 0 |
|  |  |  | Etanercept | 0 |
|  |  |  | Ustekinumab | 1 |
|  |  |  | Vedolizumab | 0 |
|  |  |  | Tofacitnib | 0 |
|  | bowel perforation = colonic perforation = peritonitis = perforation | Intestinal perforation | Infliximab | 1 |
|  |  | Large intestine perforation | Adalimumab | 4 |
|  |  | Peritonitis | Certolizumab | 0 |
|  |  | Perforation | Golimumab | 0 |
|  |  |  | Etanercept | 0 |
|  |  |  | Ustekinumab | 1 |
|  |  |  | Vedolizumab | 1 |
|  |  |  | Tofacitnib | 0 |
| **Musculoskeletal and connective tissue disorders** | Joint pains = joint pain | Arthralgia | Infliximab | 0 |
|  |  |  | Adalimumab | 1 |
|  |  |  | Certolizumab | 0 |
|  |  |  | Golimumab | 0 |
|  |  |  | Etanercept | 0 |
|  |  |  | Ustekinumab | 2 |
|  |  |  | Vedolizumab | 0 |
|  |  |  | Tofacitnib | 0 |
|  | hip fracture | Hip fracture | Infliximab | 0 |
|  |  |  | Adalimumab | 0 |
|  |  |  | Certolizumab | 0 |
|  |  |  | Golimumab | 0 |
|  |  |  | Etanercept | 0 |
|  |  |  | Ustekinumab | 0 |
|  |  |  | Vedolizumab | 1 |
|  |  |  | Tofacitnib | 0 |
|  | tenosynovitis of his right hand | Tenosynovitis | Infliximab | 0 |
|  |  |  | Adalimumab | 0 |
|  |  |  | Certolizumab | 0 |
|  |  |  | Golimumab | 0 |
|  |  |  | Etanercept | 0 |
|  |  |  | Ustekinumab | 0 |
|  |  |  | Vedolizumab | 1 |
|  |  |  | Tofacitnib | 0 |
|  | rhabdomyolysis | Rhabdomyolysis | Infliximab | 0 |
|  |  |  | Adalimumab | 1 |
|  |  |  | Certolizumab | 0 |
|  |  |  | Golimumab | 0 |
|  |  |  | Etanercept | 0 |
|  |  |  | Ustekinumab | 0 |
|  |  |  | Vedolizumab | 0 |
|  |  |  | Tofacitnib | 0 |
|  | Spasm | Muscle spasms | Infliximab | 0 |
|  |  |  | Adalimumab | 1 |
|  |  |  | Certolizumab | 0 |
|  |  |  | Golimumab | 0 |
|  |  |  | Etanercept | 0 |
|  |  |  | Ustekinumab | 0 |
|  |  |  | Vedolizumab | 0 |
|  |  |  | Tofacitnib | 0 |
|  | trismus | Trismus | Infliximab | 1 |
|  |  |  | Adalimumab | 0 |
|  |  |  | Certolizumab | 0 |
|  |  |  | Golimumab | 0 |
|  |  |  | Etanercept | 0 |
|  |  |  | Ustekinumab | 0 |
|  |  |  | Vedolizumab | 0 |
|  |  |  | Tofacitnib | 0 |
| **Gastrointestinal disorders** | Nausea and vomiting = vomiting = nausea/vomiting | Nausea | Infliximab | 6 |
|  |  | Vomiting | Adalimumab | 10 |
|  |  |  | Certolizumab | 2 |
|  |  |  | Golimumab | 0 |
|  |  |  | Etanercept | 0 |
|  |  |  | Ustekinumab | 8 |
|  |  |  | Vedolizumab | 6 |
|  |  |  | Tofacitnib | 0 |
|  | bloating = abdominal distention | Abdominal distension | Infliximab | 2 |
|  |  |  | Adalimumab | 0 |
|  |  |  | Certolizumab | 0 |
|  |  |  | Golimumab | 0 |
|  |  |  | Etanercept | 0 |
|  |  |  | Ustekinumab | 0 |
|  |  |  | Vedolizumab | 0 |
|  |  |  | Tofacitnib | 0 |
|  | internal hernia | Internal hernia | Infliximab | 0 |
|  |  |  | Adalimumab | 0 |
|  |  |  | Certolizumab | 0 |
|  |  |  | Golimumab | 0 |
|  |  |  | Etanercept | 0 |
|  |  |  | Ustekinumab | 1 |
|  |  |  | Vedolizumab | 0 |
|  |  |  | Tofacitnib | 0 |
|  | GI bleed = melena = acute GI bleeding = UGIB = upper GI bleed = GI bleeding | Gastrointestinal haemorrhage | Infliximab | 2 |
|  |  |  | Adalimumab | 1 |
|  |  |  | Certolizumab | 0 |
|  |  |  | Golimumab | 0 |
|  |  |  | Etanercept | 0 |
|  |  |  | Ustekinumab | 3 |
|  |  |  | Vedolizumab | 0 |
|  |  |  | Tofacitnib | 0 |
|  | intussusception | Intussusception | Infliximab | 0 |
|  |  |  | Adalimumab | 1 |
|  |  |  | Certolizumab | 0 |
|  |  |  | Golimumab | 0 |
|  |  |  | Etanercept | 0 |
|  |  |  | Ustekinumab | 0 |
|  |  |  | Vedolizumab | 0 |
|  |  |  | Tofacitnib | 0 |
|  | pancreatitis | Pancreatitis | Infliximab | 0 |
|  |  |  | Adalimumab | 1 |
|  |  |  | Certolizumab | 0 |
|  |  |  | Golimumab | 0 |
|  |  |  | Etanercept | 0 |
|  |  |  | Ustekinumab | 0 |
|  |  |  | Vedolizumab | 0 |
|  |  |  | Tofacitnib | 0 |
| **General disorders and administration site conditions** | Fever = high fever | Pyrexia | Infliximab | 8 |
|  |  |  | Adalimumab | 2 |
|  |  |  | Certolizumab | 0 |
|  |  |  | Golimumab | 0 |
|  |  |  | Etanercept | 0 |
|  |  |  | Ustekinumab | 1 |
|  |  |  | Vedolizumab | 3 |
|  |  |  | Tofacitnib | 0 |
|  | Fatigue = tired = fatigued | Fatigue | Infliximab | 4 |
|  |  |  | Adalimumab | 1 |
|  |  |  | Certolizumab | 1 |
|  |  |  | Golimumab | 0 |
|  |  |  | Etanercept | 0 |
|  |  |  | Ustekinumab | 0 |
|  |  |  | Vedolizumab | 2 |
|  |  |  | Tofacitnib | 0 |
|  | edema = swelling = neck, and painful swelling | Oedema | Infliximab | 0 |
|  |  | Swelling | Adalimumab | 1 |
|  |  |  | Certolizumab | 0 |
|  |  |  | Golimumab | 0 |
|  |  |  | Etanercept | 0 |
|  |  |  | Ustekinumab | 0 |
|  |  |  | Vedolizumab | 2 |
|  |  |  | Tofacitnib | 0 |
|  | weakness | Asthenia | Infliximab | 0 |
|  |  |  | Adalimumab | 0 |
|  |  |  | Certolizumab | 1 |
|  |  |  | Golimumab | 0 |
|  |  |  | Etanercept | 0 |
|  |  |  | Ustekinumab | 0 |
|  |  |  | Vedolizumab | 1 |
|  |  |  | Tofacitnib | 0 |
|  | rigors = chills | Chills | Infliximab | 1 |
|  |  |  | Adalimumab | 2 |
|  |  |  | Certolizumab | 0 |
|  |  |  | Golimumab | 0 |
|  |  |  | Etanercept | 0 |
|  |  |  | Ustekinumab | 0 |
|  |  |  | Vedolizumab | 0 |
|  |  |  | Tofacitnib | 0 |
|  | mass | Mass | Infliximab | 0 |
|  |  |  | Adalimumab | 0 |
|  |  |  | Certolizumab | 0 |
|  |  |  | Golimumab | 0 |
|  |  |  | Etanercept | 0 |
|  |  |  | Ustekinumab | 0 |
|  |  |  | Vedolizumab | 1 |
|  |  |  | Tofacitnib | 0 |
| **Blood and lymphatic system disorders** | Thrombocytopenia | Thrombocytopenia | Infliximab | 1 |
|  |  |  | Adalimumab | 0 |
|  |  |  | Certolizumab | 0 |
|  |  |  | Golimumab | 0 |
|  |  |  | Etanercept | 0 |
|  |  |  | Ustekinumab | 0 |
|  |  |  | Vedolizumab | 0 |
|  |  |  | Tofacitnib | 0 |
|  | lymphadenopathy | Lymphadenopathy | Infliximab | 0 |
|  |  |  | Adalimumab | 0 |
|  |  |  | Certolizumab | 0 |
|  |  |  | Golimumab | 0 |
|  |  |  | Etanercept | 0 |
|  |  |  | Ustekinumab | 0 |
|  |  |  | Vedolizumab | 2 |
|  |  |  | Tofacitnib | 0 |
|  | leukocytosis | Leukocytosis | Infliximab | 0 |
|  |  |  | Adalimumab | 0 |
|  |  |  | Certolizumab | 0 |
|  |  |  | Golimumab | 0 |
|  |  |  | Etanercept | 0 |
|  |  |  | Ustekinumab | 1 |
|  |  |  | Vedolizumab | 0 |
|  |  |  | Tofacitnib | 0 |
| **Psychiatric disorders** | confusion | Confusional state | Infliximab | 1 |
|  |  |  | Adalimumab | 0 |
|  |  |  | Certolizumab | 0 |
|  |  |  | Golimumab | 0 |
|  |  |  | Etanercept | 0 |
|  |  |  | Ustekinumab | 0 |
|  |  |  | Vedolizumab | 0 |
|  |  |  | Tofacitnib | 0 |
|  | psychosis | Psychotic disorder | Infliximab | 0 |
|  |  |  | Adalimumab | 0 |
|  |  |  | Certolizumab | 0 |
|  |  |  | Golimumab | 0 |
|  |  |  | Etanercept | 0 |
|  |  |  | Ustekinumab | 0 |
|  |  |  | Vedolizumab | 1 |
|  |  |  | Tofacitnib | 0 |
| **Endocrine disorders** | Adrenal insufficiency | Adrenal insufficiency | Infliximab | 1 |
|  |  |  | Adalimumab | 0 |
|  |  |  | Certolizumab | 0 |
|  |  |  | Golimumab | 0 |
|  |  |  | Etanercept | 0 |
|  |  |  | Ustekinumab | 0 |
|  |  |  | Vedolizumab | 0 |
|  |  |  | Tofacitnib | 0 |
|  | hyperthyroidism | Hyperthyroidism | Infliximab | 0 |
|  |  |  | Adalimumab | 0 |
|  |  |  | Certolizumab | 0 |
|  |  |  | Golimumab | 0 |
|  |  |  | Etanercept | 0 |
|  |  |  | Ustekinumab | 0 |
|  |  |  | Vedolizumab | 1 |
|  |  |  | Tofacitnib | 0 |
| **Cardiac disorders** | tachycardia | Tachycardia | Infliximab | 1 |
|  |  |  | Adalimumab | 0 |
|  |  |  | Certolizumab | 0 |
|  |  |  | Golimumab | 0 |
|  |  |  | Etanercept | 0 |
|  |  |  | Ustekinumab | 0 |
|  |  |  | Vedolizumab | 0 |
|  |  |  | Tofacitnib | 0 |
|  | cardiomyopathy | Cardiomyopathy | Infliximab | 0 |
|  |  |  | Adalimumab | 1 |
|  |  |  | Certolizumab | 0 |
|  |  |  | Golimumab | 0 |
|  |  |  | Etanercept | 0 |
|  |  |  | Ustekinumab | 0 |
|  |  |  | Vedolizumab | 0 |
|  |  |  | Tofacitnib | 0 |
|  | bradycardia | Bradycardia | Infliximab | 0 |
|  |  |  | Adalimumab | 0 |
|  |  |  | Certolizumab | 0 |
|  |  |  | Golimumab | 0 |
|  |  |  | Etanercept | 0 |
|  |  |  | Ustekinumab | 1 |
|  |  |  | Vedolizumab | 0 |
|  |  |  | Tofacitnib | 0 |
|  | myopericarditis | Myocarditis | Infliximab | 1 |
|  |  |  | Adalimumab | 0 |
|  |  |  | Certolizumab | 0 |
|  |  |  | Golimumab | 0 |
|  |  |  | Etanercept | 0 |
|  |  |  | Ustekinumab | 0 |
|  |  |  | Vedolizumab | 0 |
|  |  |  | Tofacitnib | 0 |
|  | arrhythmia | Arrhythmia | Infliximab | 1 |
|  |  |  | Adalimumab | 0 |
|  |  |  | Certolizumab | 0 |
|  |  |  | Golimumab | 0 |
|  |  |  | Etanercept | 0 |
|  |  |  | Ustekinumab | 0 |
|  |  |  | Vedolizumab | 0 |
|  |  |  | Tofacitnib | 0 |
|  | chest pain | Chest pain | Infliximab | 0 |
|  |  |  | Adalimumab | 1 |
|  |  |  | Certolizumab | 0 |
|  |  |  | Golimumab | 0 |
|  |  |  | Etanercept | 0 |
|  |  |  | Ustekinumab | 0 |
|  |  |  | Vedolizumab | 0 |
|  |  |  | Tofacitnib | 0 |
| **Respiratory, thoracic and mediastinal disorders** | Pleural effusion = pleural effusions | Pleural effusion | Infliximab | 2 |
|  |  |  | Adalimumab | 0 |
|  |  |  | Certolizumab | 0 |
|  |  |  | Golimumab | 0 |
|  |  |  | Etanercept | 0 |
|  |  |  | Ustekinumab | 0 |
|  |  |  | Vedolizumab | 0 |
|  |  |  | Tofacitnib | 0 |
|  | SOB = shortness of breath | Dyspnoea | Infliximab | 1 |
|  |  |  | Adalimumab | 0 |
|  |  |  | Certolizumab | 0 |
|  |  |  | Golimumab | 0 |
|  |  |  | Etanercept | 0 |
|  |  |  | Ustekinumab | 0 |
|  |  |  | Vedolizumab | 1 |
|  |  |  | Tofacitnib | 0 |
|  | stridor | Stridor | Infliximab | 0 |
|  |  |  | Adalimumab | 0 |
|  |  |  | Certolizumab | 0 |
|  |  |  | Golimumab | 0 |
|  |  |  | Etanercept | 0 |
|  |  |  | Ustekinumab | 0 |
|  |  |  | Vedolizumab | 1 |
|  |  |  | Tofacitnib | 0 |
|  | coughing | Cough | Infliximab | 0 |
|  |  |  | Adalimumab | 0 |
|  |  |  | Certolizumab | 0 |
|  |  |  | Golimumab | 0 |
|  |  |  | Etanercept | 0 |
|  |  |  | Ustekinumab | 0 |
|  |  |  | Vedolizumab | 1 |
|  |  |  | Tofacitnib | 0 |
| **Renal and urinary disorders** | Minimal change nephrotic syndrome | Nephrotic syndrome | Infliximab | 0 |
|  |  |  | Adalimumab | 0 |
|  |  |  | Certolizumab | 0 |
|  |  |  | Golimumab | 0 |
|  |  |  | Etanercept | 0 |
|  |  |  | Ustekinumab | 0 |
|  |  |  | Vedolizumab | 1 |
|  |  |  | Tofacitnib | 0 |
|  | ARF = acute renal failure | Acute kidney injury | Infliximab | 0 |
|  |  |  | Adalimumab | 1 |
|  |  |  | Certolizumab | 0 |
|  |  |  | Golimumab | 0 |
|  |  |  | Etanercept | 0 |
|  |  |  | Ustekinumab | 0 |
|  |  |  | Vedolizumab | 0 |
|  |  |  | Tofacitnib | 0 |
|  | ATN = acute tubular necrosis | Renal tubular necrosis | Infliximab | 0 |
|  |  |  | Adalimumab | 1 |
|  |  |  | Certolizumab | 0 |
|  |  |  | Golimumab | 0 |
|  |  |  | Etanercept | 0 |
|  |  |  | Ustekinumab | 0 |
|  |  |  | Vedolizumab | 0 |
|  |  |  | Tofacitnib | 0 |
| **Nervous system disorders** | complex migraine = headaches | Migraine | Infliximab | 0 |
|  |  | Headache | Adalimumab | 3 |
|  |  |  | Certolizumab | 0 |
|  |  |  | Golimumab | 0 |
|  |  |  | Etanercept | 0 |
|  |  |  | Ustekinumab | 0 |
|  |  |  | Vedolizumab | 0 |
|  |  |  | Tofacitnib | 0 |
|  | Encephalitis seizure = seizures | Encephalitis | Infliximab | 1 |
|  |  | Seizure | Adalimumab | 0 |
|  |  |  | Certolizumab | 0 |
|  |  |  | Golimumab | 0 |
|  |  |  | Etanercept | 0 |
|  |  |  | Ustekinumab | 0 |
|  |  |  | Vedolizumab | 1 |
|  |  |  | Tofacitnib | 0 |
|  | metabolic encephalopathy | Metabolic encephalopathy | Infliximab | 0 |
|  |  |  | Adalimumab | 1 |
|  |  |  | Certolizumab | 0 |
|  |  |  | Golimumab | 0 |
|  |  |  | Etanercept | 0 |
|  |  |  | Ustekinumab | 0 |
|  |  |  | Vedolizumab | 0 |
|  |  |  | Tofacitnib | 0 |
|  | stroke | Cerebrovascular accident | Infliximab | 1 |
|  |  |  | Adalimumab | 0 |
|  |  |  | Certolizumab | 0 |
|  |  |  | Golimumab | 0 |
|  |  |  | Etanercept | 0 |
|  |  |  | Ustekinumab | 1 |
|  |  |  | Vedolizumab | 1 |
|  |  |  | Tofacitnib | 0 |
|  | syncope = near syncope | Syncope | Infliximab | 0 |
|  |  | Presyncope | Adalimumab | 1 |
|  |  |  | Certolizumab | 0 |
|  |  |  | Golimumab | 0 |
|  |  |  | Etanercept | 0 |
|  |  |  | Ustekinumab | 1 |
|  |  |  | Vedolizumab | 0 |
|  |  |  | Tofacitnib | 0 |
| **Eye disorders** | uveitis | Uveitis | Infliximab | 0 |
|  |  |  | Adalimumab | 1 |
|  |  |  | Certolizumab | 0 |
|  |  |  | Golimumab | 0 |
|  |  |  | Etanercept | 0 |
|  |  |  | Ustekinumab | 0 |
|  |  |  | Vedolizumab | 0 |
|  |  |  | Tofacitnib | 0 |
| **Investigations** | Elevated liver enzymes = rising LFTs = hepatitis | Hepatic enzyme increased | Infliximab | 3 |
|  |  | Hepatitis | Adalimumab | 0 |
|  |  |  | Certolizumab | 0 |
|  |  |  | Golimumab | 0 |
|  |  |  | Etanercept | 0 |
|  |  |  | Ustekinumab | 0 |
|  |  |  | Vedolizumab | 0 |
|  |  |  | Tofacitnib | 0 |
| **Hepatobiliary disorders** | cholangitis | Cholangitis | Infliximab | 1 |
|  |  |  | Adalimumab | 0 |
|  |  |  | Certolizumab | 0 |
|  |  |  | Golimumab | 0 |
|  |  |  | Etanercept | 0 |
|  |  |  | Ustekinumab | 0 |
|  |  |  | Vedolizumab | 0 |
|  |  |  | Tofacitnib | 0 |
|  | choledocholithiasis | Bile duct stone | Infliximab | 0 |
|  |  |  | Adalimumab | 0 |
|  |  |  | Certolizumab | 0 |
|  |  |  | Golimumab | 0 |
|  |  |  | Etanercept | 0 |
|  |  |  | Ustekinumab | 1 |
|  |  |  | Vedolizumab | 0 |
|  |  |  | Tofacitnib | 0 |
| **Injury, poisoning and procedural complications** | hepatic artery injury | Arterial injury | Infliximab | 1 |
|  |  |  | Adalimumab | 0 |
|  |  |  | Certolizumab | 0 |
|  |  |  | Golimumab | 0 |
|  |  |  | Etanercept | 0 |
|  |  |  | Ustekinumab | 0 |
|  |  |  | Vedolizumab | 0 |
|  |  |  | Tofacitnib | 0 |
|  | anastomotic leak | Anastomotic leak | Infliximab | 1 |
|  |  |  | Adalimumab | 0 |
|  |  |  | Certolizumab | 0 |
|  |  |  | Golimumab | 0 |
|  |  |  | Etanercept | 0 |
|  |  |  | Ustekinumab | 0 |
|  |  |  | Vedolizumab | 0 |
|  |  |  | Tofacitnib | 0 |
| **Metabolism and nutrition disorders** | weight loss = protein calorie malnutrition = poor appetite = inability to tolerate PO = difficulty tolerating PO = failure to thrive = malnutrition | Weight decreased | Infliximab | 3 |
|  |  | Malnutrition | Adalimumab | 0 |
|  |  | Decreased appetite | Certolizumab | 1 |
|  |  | Failure to thrive | Golimumab | 0 |
|  |  |  | Etanercept | 0 |
|  |  |  | Ustekinumab | 4 |
|  |  |  | Vedolizumab | 4 |
|  |  |  | Tofacitnib | 0 |
|  | dehydration | Dehydration | Infliximab | 0 |
|  |  |  | Adalimumab | 3 |
|  |  |  | Certolizumab | 0 |
|  |  |  | Golimumab | 0 |
|  |  |  | Etanercept | 0 |
|  |  |  | Ustekinumab | 1 |
|  |  |  | Vedolizumab | 2 |
|  |  |  | Tofacitnib | 0 |
|  | hypokalemia | Hypokalaemia | Infliximab | 0 |
|  |  |  | Adalimumab | 0 |
|  |  |  | Certolizumab | 0 |
|  |  |  | Golimumab | 0 |
|  |  |  | Etanercept | 0 |
|  |  |  | Ustekinumab | 1 |
|  |  |  | Vedolizumab | 0 |
|  |  |  | Tofacitnib | 0 |
|  | electrolyte abnormalities | Blood electrolytes abnormal | Infliximab | 0 |
|  |  |  | Adalimumab | 0 |
|  |  |  | Certolizumab | 0 |
|  |  |  | Golimumab | 0 |
|  |  |  | Etanercept | 0 |
|  |  |  | Ustekinumab | 0 |
|  |  |  | Vedolizumab | 1 |
|  |  |  | Tofacitnib | 0 |
| **Vascular disorders** | hypotension = hypovolemia | Hypotension | Infliximab | 2 |
|  |  | Hypovolaemia | Adalimumab | 0 |
|  |  |  | Certolizumab | 0 |
|  |  |  | Golimumab | 0 |
|  |  |  | Etanercept | 0 |
|  |  |  | Ustekinumab | 0 |
|  |  |  | Vedolizumab | 0 |
|  |  |  | Tofacitnib | 0 |
|  | hemorrhage shock | Shock haemorrhagic | Infliximab | 0 |
|  |  |  | Adalimumab | 0 |
|  |  |  | Certolizumab | 0 |
|  |  |  | Golimumab | 0 |
|  |  |  | Etanercept | 0 |
|  |  |  | Ustekinumab | 1 |
|  |  |  | Vedolizumab | 0 |
|  |  |  | Tofacitnib | 0 |
|  | DVT = deep vein thrombosis = PE = pulmonary embolism = DVTs | Deep vein thrombosis | Infliximab | 3 |
|  |  | Pulmonary embolism | Adalimumab | 1 |
|  |  |  | Certolizumab | 0 |
|  |  |  | Golimumab | 0 |
|  |  |  | Etanercept | 0 |
|  |  |  | Ustekinumab | 1 |
|  |  |  | Vedolizumab | 0 |
|  |  |  | Tofacitnib | 0 |
| **Neoplasms benign, malignant and unspecified (incl cysts and polyps)** | sarcoma | Sarcoma | Infliximab | 1 |
|  |  |  | Adalimumab | 0 |
|  |  |  | Certolizumab | 0 |
|  |  |  | Golimumab | 0 |
|  |  |  | Etanercept | 0 |
|  |  |  | Ustekinumab | 0 |
|  |  |  | Vedolizumab | 0 |
|  |  |  | Tofacitnib | 0 |
| **Skin and subcutaneous tissue disorders** | rash | Rash | Infliximab | 1 |
|  |  |  | Adalimumab | 1 |
|  |  |  | Certolizumab | 0 |
|  |  |  | Golimumab | 0 |
|  |  |  | Etanercept | 0 |
|  |  |  | Ustekinumab | 1 |
|  |  |  | Vedolizumab | 0 |
|  |  |  | Tofacitnib | 0 |
|  | pyoderma gangrenosum | Pyoderma gangrenosum | Infliximab | 0 |
|  |  |  | Adalimumab | 0 |
|  |  |  | Certolizumab | 0 |
|  |  |  | Golimumab | 0 |
|  |  |  | Etanercept | 0 |
|  |  |  | Ustekinumab | 0 |
|  |  |  | Vedolizumab | 1 |
|  |  |  | Tofacitnib | 0 |

### All Models Tested and their Performance

Supplemental Table 4. All models tested and their performance. Bolded text corresponds to highest performing model.

| Task | Model | Accuracy (%) | Macro F1 (%) | TP | TN | FP | FN |
| --- | --- | --- | --- | --- | --- | --- | --- |
| Medication before hospitalization relations | **H-UCSF-BERT** | **88** | **62** | 63 | 1989 | 173 | 105 |
|  | Logistic Regression | 72 | 41 | 42 | 1653 | 509 | 126 |
|  | XGBoost | 73 | 51 | 49 | 1672 | 490 | 119 |
|  | CNN | 74 | 49 | 51 | 1682 | 480 | 117 |
|  | BiLSTM | 73 | 48 | 51 | 1670 | 492 | 117 |
| Hospitalization for SAE relations | H-UCSF-BERT | 96 | 62 | 79 | 9603 | 421 | 16 |
|  | **H-UCSF-BERT + only nearby SAEs** | **92** | **68** | 34 | 1078 | 41 | 61 |
|  | KNNeighbors + only nearby SAEs | 88 | 53 | 11 | 1064 | 55 | 84 |
|  | Decision Tree + only nearby SAEs | 86 | 55 | 18 | 1038 | 81 | 77 |
|  | CNN + only nearby AEs | 92 | 47 | 0 | 1119 | 0 | 95 |
|  | BiLSTM + only nearby SAEs | 93 | 48 | 7 | 1091 | 28 | 88 |
|  | Decision trees (BoW) + only nearby SAEs | 88 | 53 | 0 | 8409 | 0 | 319 |
| Medication before hospitalization for adverse event relation (triples) | **H-UCSF-BERT + only nearby SAEs** | **91** | **61** | **141** | **7790** | **619** | **178** |
|  | CNN + only nearby AEs | 94 | 49 | 11 | 8013 | 396 | 308 |
|  | BiLSTM + only nearby AEs | 95 | 50 | 11 | 7953 | 456 | 308 |
|  | BiLSTM + attention + only nearby AEs | 94 | 49 | 0 | 8409 | 0 | 319 |
|  | AutoML: NeuralNetFastAI_BAG_L1 + only nearby AEs | 84 | 58 | 25 | 7362 | 1047 | 294 |
